# Factors influencing the sustainability of homestead vegetable production intervention in Rufiji, Tanzania: A cross-sectional mixed methods study

**DOI:** 10.1101/2022.05.06.22274693

**Authors:** Killian N Mlalama, Athanasia Matemu, Ephraim Kosia, Chelsey R Canavan, Alexandra L Bellows, Mia Blakstad, Ramadhani A Noor, Joyce Kinabo, Wafaie W Fawzi, Honorati Masanja, Dominic Mosha

## Abstract

**Background:** There is growing evidence that home vegetable gardening interventions improve food security and nutrition outcomes at the family level. This study assessed factors influencing the sustainability of homestead vegetable production intervention in Rufiji district, Tanzania, one year after the cessation of external support.

**Methods:** This was a cross-sectional study using both quantitative and qualitative data collection methods. A total of 247 randomly selected women from households who participated in the homestead vegetable intervention were interviewed using a structured questionnaire. The study held four focus group discussions with women from households that participated in the intervention, and four In-Depth interviews with two extension workers, one community health worker, and one agriculture district officer. Multiple logistic regression for quantitative data and thematic analysis for qualitative data was conducted.

**Results:** About 20.24% (50/247) of households sustained homestead vegetable production for one year after the intervention phased out. Lack of seeds (adjusted OR=1.26: CI=0.39-0.89) and either manure or fertilizers (adjusted OR=1.69: CI =1.08-2.63) were significant factors influencing the sustainability of homesteads vegetable production. In the Focus Group discussions (FGDs) and In-Depth Interview (IDIs), all participating women and extension workers reported high cost of water, destruction from free-grazing animals, agriculture pests and diseases, poor soil fertility, shortage of seeds, and lack of capital affected homestead vegetable production sustainability.

**Conclusion:** Existing individual, community, and system challenges influence the sustainability of external-funded agriculture and nutrition interventions. The study findings underscore the importance of community authorities, scientists, and policymakers in having a well-thought sustainability plan in all promising external-funded interventions.

## Introduction

The global statistics report that about 2 billion persons globally experience micronutrient deficiencies, and at least 800 million suffer from energy deficiencies (1). Sub-Saharan Africa is undergoing the burden of micronutrient and nutrition deficiency with reported high ranks of malnutrition and increasing rate of obesity, overweight, underweight, wasting and stunting as diet-related diseases (2,3). For example, in East Africa, the prevalence of stunting for under-five children is high as 34% (4), with 32.1% in Tanzania(5). Rufiji district in the Coastal region has 35% of under-five stunting prevalence, which is higher than that of Tanzania country as a whole (6). Poverty associated factors like food insecurity, floods and drought are the most stated drivers of malnutrition (7). Different strategies to reduce the undernutrition burden are ongoing through integrating government, donor agencies, and development organizations to support nutrition-sensitive interventions.

Vegetable consumption is highly recognized to control malnutrition as it provides different nutrients to combat mineral and vitamin deficiencies, denoted as “hidden hunger” (8). This underscores the importance of the agriculture sector, one of the components of nutrition-sensitive interventions to improving nutrition, specifically through integrated agriculture and nutrition programmes such as homestead vegetable gardening (9). Home vegetable production has become an effective strategy to reduce undernutrition and micronutrient deficiency at the household level in many developing countries(10).

Despite the well-known benefits of homestead vegetable production (11–13), uptake of vegetable gardening is still low in Sub Saharan African countries, for example Southern district of Botswana(14). This situation, eventually leads to low vegetable consumption as recommended by World Health Organization (15).

Besides, there is inadequate evidence of the studies that have investigated the sustainability of agriculture nutrition-sensitive intervention projects implemented in different parts of the world, including Tanzania. The only few studies conducted on post-intervention sustainability of homestead vegetable gardening includes the one conducted in Dakar, Senegal which found only 5% of the participants sustained growing vegetable gardens 18 months after the project phased-out (16). In another study conducted in a rural village in South Africa, 39% of the participants kept growing vegetable gardens ten years after cessation of homestead vegetable production project (17). Limitations reported hindering sustainability of homestead vegetable donor-funded project in developing countries includes space constraints, poverty, lack of fencing, shortage of water, animals eating crops and lack of support services (16–18). Bearing in mind that most of the interventions are donor-funded, and agencies supporting usually withdraw from the communities after the programs phase-out, it was important to investigate this research gap.

Therefore, this study intends to understand factors influencing the sustainability of homestead vegetable production one-year after the phase-out of the agriculture nutrition-sensitive intervention in Rufiji, Tanzania.

## Materials and methods

### Study area

This study was conducted in Rufiji district, located in eastern Tanzania 7.47° to 8.03° south latitude and 38.62° to 39.17° east longitude (19). The district demographic characteristics are representative of rural Tanzania, most adults are subsistence farmers who cultivate cashew nuts, sesame, coconuts, and staples like maize, rice, cassava and millet (20). The prevalence of stunting and anemia among children under-five years in Rufiji is 35% and 61%, respectively (6).

### Description of the intervention

The Homestead Agriculture and Nutrition (HANU) intervention was a cluster randomized controlled trial (RCT) conducted in 10 villages, 5 intervention and the other 5 control villages. The intervention operated for three years, from August 2016 to December 2019. The aim of the study was to determine the effects of integrated agriculture nutrition-sensitive interventions on the health and nutrition of children and women in rural Tanzania while exploring the incorporation of existing cadres of agricultural extension workers and community health workers as a sustainable workforce for providing basic health and nutrition education in rural communities.

Participants in the intervention arm received three main intervention packages: (i) agriculture inputs such as seeds, watering cans, manure, fertilizers, and training to promote homestead food production and increase food diversity, (ii) nutrition counselling, including prevention and management of child malnutrition, and (iii) a health-focused intervention including information on micronutrient supplementation, integrated management of child illness, and safe water, sanitation and hygiene practices. The extension and community health workers provided these training and educational messages through household visits and farm field school training. A detailed description of the design, sampling procedures and inclusion criteria have been presented in detail elsewhere (21).

### Study design

This was a cross-sectional study design using a mixed-methods sequential explanatory data collection approaches, where quantitative data analysis results led to development of themes for qualitative survey (22). The study was conducted from April to June 2021, one year after the phase-out of HANU trial. The population for this study was women who previously participated in the Homestead Agriculture and Nutrition (HANU) trial in 5 randomly selected intervention villages.

Participants’ source list was acquired from the HANU trial database, where all 500 participants from five intervention villages were subjected to simple random sampling. A proportion-based representation of intervention households per village was considered when sampling to obtain a representative sample size from each village. Household replacement was considered after missing the participant in two consecutive visits. A randomized household list for replacement in each intervention village was also generated, and a standard orderly sequence was applied to guide selecting households from the list for replacement. This involved collecting data at one point from 247 randomly selected households. A simple random sampling technique was used for the quantitative survey to select women who participated in the HANU trial under the intervention arm.

For qualitative survey, purposive sampling was performed to achieve uniform representation of the four Focus Group Discussions (FGDs) of women from five villages that received the HANU intervention; two FGDs involved women actively practicing home gardening to date and two FGDs involved women not practicing home gardening since the end of the HANU trial. We conducted four In-Depth Interviews (IDIs) with two extension workers, one community health worker, and one agriculture district officer.

### Sample size

The sample size for the quantitative survey was pre-determined by the number of participants in the trial under the intervention arm (500 women) and the logically feasible time frame of three months. The number of participants in the intervention trial arm who could be enrolled was estimated before (as 250) to be sufficient to determine sustainability status of the intervention, but no formal sample size calculation was performed.

### Data collection

Quantitative data collection was done by trained research assistants using closed-ended structured questionnaires administered in electronic devices (tablets). The collected data was then uploaded and stored to Ifakara Health Institute(IHI) server in Dar es salaam, Tanzania for safe data management.

Open-ended questions were used to generate information from a total of four FGDs and four IDIs. A digital voice recorder was used to capture and store voice before verbatim transcription. FGDs and IDIs explored participants’ perceptions of potential strengths, weaknesses, opportunities, and threats that may influence the sustainability of vegetable gardening in the absence of external financial and technical support. Each FGD had 9 to 12 participants. FGDs and IDIs were conducted using a topic guide that had been pilot-tested. Two experienced qualitative researchers conducted FGD and IDI in Swahili language and in a private place to ensure confidentiality. All the FGDs and IDIs were audio-recorded and transcribed verbatim and later translated into English.

### Data management and analysis

Quantitative data were cleaned and analyzed by using STATA version 15. Numerical variables were summarized using means and standard deviations. Categorical variables were summarized using frequencies and percentages. The effect of demographic characteristics and agriculture resources on primary endpoint of the study (sustaining gardening) was assessed by bivariate analysis.

To get a deeper understanding of the data on how different variables impact sustainability of homestead vegetable gardening, logistic regression models were used to estimate the odds ratio (OR) for the association between active practising vegetable gardening status and potential agriculture resources. Explanatory variables were included in the multivariate analysis using the forward regression approach if the variable had a p-value <0.2 in the bivariate analysis or if there was a strong knowledge supporting the effect of a particular independent variable on the primary outcome, such factors were included in the multivariable logistic regression regardless of their p-values being >0.2.

For FGDs and IDIs, thematic analysis was done with the aid of NVivo 12 pro qualitative analysis software (23). Transcripts were imported into NVivo, and two experienced social scientists did the coding. The list of codes and main theme frequencies were summarized. Only themes that answered the specific study objective were identified during the thematic analysis. Furthermore, themes from all five study villages including FGDs and IDIs were combined after thematic analysis due to the similarity of their results.

### Ethical considerations

Ethical clearance to conduct this study was approved by the Institutional Review Board of Ifakara Health Institute (IHI) Dar es Salaam, Tanzania. Ethical approval number IHI/IRB/No:13-2021 was granted. The Principal Investigator assured that the study was conducted in full compliance to the ethical principles of the “Declaration of Helsinki” and all laws and regulations of the United Republic of Tanzania. Written informed consent was obtained from all study participants. Confidentiality and anonymity of participants’ data were highly maintained.

## Results

A total of 247 women were recruited and interviewed in the quantitative survey with the mean (sd) of age 36 (8.3) years; 85.0% were married or cohabiting, 35.2% did not attend formal education, and 73.7% lived in their own house or residence. The mean family size was 7.1 (2.6) persons (Table 1).

**Table 1.** Social-demographic characteristics of study participants.

| **Variable** | **Frequency**<br>**(n=247)** | **Percentage**<br>**(%)** |
| --- | --- | --- |
| **Household size, mean (SD)** | 7.1 (2.6) |  |
| **Level of Education** |  |  |
| No formal education | 87 | 35.22 |
| Primary school education | 141 | 57.09 |
| Secondary school education | 19 | 7.69 |
| **Age of respondent (years)** |  |  |
| Average age, mean (SD) | 36.0 (8.3) |  |
| **Marital status** |  |  |
| Married | 210 | 85.02 |
| Single | 15 | 6.07 |
| Widowed | 5 | 2.02 |
| Divorced | 17 | 6.88 |
| **Home ownership** |  |  |
| Rent a house | 18 | 7.29 |
| Stay with relatives | 45 | 18.22 |
| Own my house | 182 | 73.68 |
| Others | 2 | 0.81 |
| **Occupation** |  |  |
| Farming | 207 | 83.81 |
| Business | 13 | 5.26 |
| Self employed | 17 | 6.88 |
| Others | 10 | 4.05 |
| **House roofing** |  |  |
| Thatched | 34 | 13.77 |
| Metal (tin or aluminium) | 210 | 85.02 |
| Dry grasses | 3 | 1.21 |

About 20.24% (50/247) participants reported actively participating in vegetable gardening one year after the nutrition-sensitive homestead agriculture intervention had phase-out. All participants with active vegetable gardens were growing cassava, sweet potato, *moringa*, and pumpkin, considered traditional vegetables whose seeds/seedlings are locally available. Less than 30% of participants with active vegetable gardens were growing amaranth, tomato, African egg-plant, and Chinese-cabbage which participants were supplied with seeds during the intervention phase (Fig 1).

**Fig 1.**
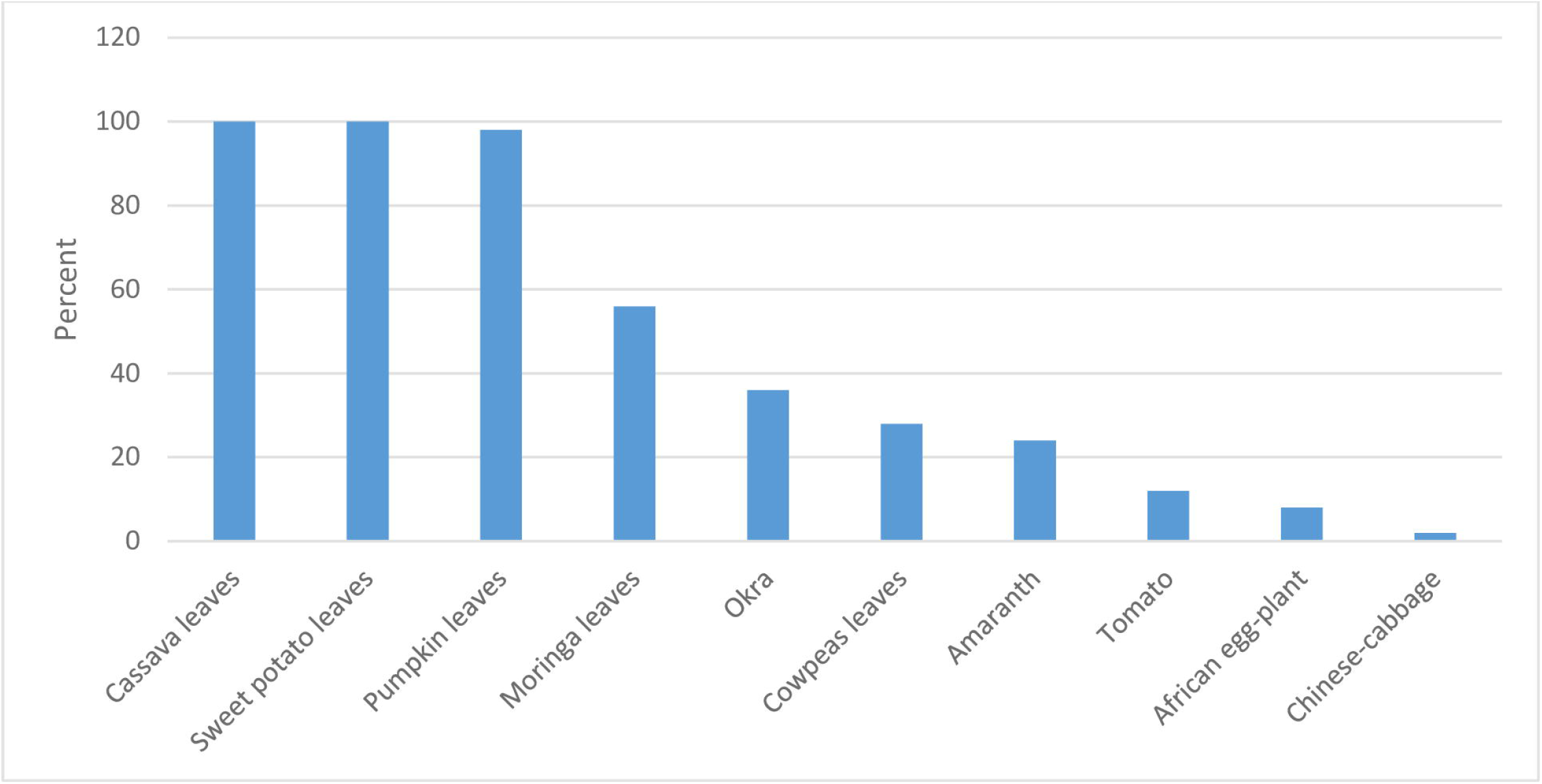
Top 10 vegetable varieties grown one year after the intervention phase-out.

In the multiple logistic regression predictive models for the sustainability of home vegetable gardens, participants in households who could buy seeds were significantly more likely to have active home vegetable gardens (adjusted OR=1.26; 95%CI= 0.39 - 0.89; p = 0.013). In addition, participants in households that did not experience a shortage of manure and fertilizers were significantly more likely to have active home vegetable gardens (adjusted OR =1.69; 95%CI= 1.08 - 2.63; p = 0.020) (Table 2).

**Table 2.** Factors associated with sustainability of home vegetable gardening intervention in Rufiji, Tanzania.

| Variable | Bivariate Analysis |  |  | Multivariate Regression Analysis |  |  |
| --- | --- | --- | --- | --- | --- | --- |
|  | COR | 95% CI | P-Value | AOR | 95% CI | P-Value |
| Water cost | 0.83 | 0.62-1.12 | 0.231 | 0.80 | 0.49-1.31 | 0.375 |
| Destruction by free animals | 0.70 | 0.50-0.97 | 0.033 | 0.80 | 0.55-1.17 | 0.256 |
| Pest and diseases | 0.98 | 0.71-1.36 | 0.921 | 0.97 | 0.65-1.44 | 0.871 |
| Water shortage | 0.91 | 0.69-1.21 | 0.531 | 1.11 | 0.71-1.73 | 0.656 |
| Lack of capital | 1.32 | 0.93-1.88 | 0.122 | 1.40 | 0.86-2.29 | 0.174 |
| Poor soils | 1.38 | 0.97-1.97 | 0.077 | 1.23 | 0.78-1.91 | 0.375 |
| Weeds | 1.02 | 0.61-1.69 | 0.950 | 0.97 | 0.55-1.7 | 0.924 |
| Seeds shortage | 1.82 | 0.63-1.07 | 0.146 | **1.26** | **0.39-0.89** | **0.013** |
| Lack of extension services | 0.94 | 0.72-1.23 | 0.677 | 0.77 | 0.53-1.11 | 0.163 |
| Cost of agriculture inputs | 0.97 | 0.75-1.25 | 0.825 | 1.07 | 0.67-1.69 | 0.773 |
| Limited family labor | 1.06 | 0.74-1.54 | 0.742 | 1.03 | 0.62-1.70 | 0.910 |
| Lack of time to work | 1.19 | 0.76-1.86 | 0.438 | 1.14 | 0.65-1.99 | 0.650 |
| Theft | 1.01 | 0.42-2.46 | 0.978 | 0.96 | 0.37-2.52 | 0.942 |
| Damage by weather | 1.07 | 0.70-1.64 | 0.745 | 1.31 | 0.72-2.38 | 0.379 |
| Shortage of manure & fertilizers | 1.62 | 1.15-2.27 | 0.006 | **1.69** | **1.08-2.63** | **0.020** |

Abbreviations: Figures in boldface are significant statistical relationship, COR, crude odds ratio, AOR, adjusted odds ratio, CI<sub>95%</sub>, 95% confidence interval.
Adjusted odds ratio for probable confounding effect-adjusted for water cost, destruction by free animals, pests and diseases, water shortage, lack of capital, poor soils, weeds, shortage of seeds, lack of extension services, cost of agriculture inputs, limited family labour, lack of time to work, theft, damage by weather, and shortage of manure and fertilizers.

In FGDs and IDIs, factors mostly reported by participants to influence the sustainability of homestead vegetable production include garden and vegetable damage caused by free-grazing animals, cost of water, pests and diseases, shortage of seeds, shortage of manure and fertilizers.

Majority of the participants were not able to build a strong fence for garden protection from free-grazing poultry and animals such as goats, chickens, and ducks. Due to the high cost of hard and strong fencing materials, most gardeners built short-term fences using coconut thatches, sesame trees, and old bed nets.

> *“…… lack of a strong fence to prevent chickens and goats. I often build temporary low cost fence using sesame trees, thatches or normal dry grasses however, chickens and goats push and penetrate to attack vegetables inside the garden. But if I were able to make a strong fence around the garden, chickens and goats would not be able to enter the garden”* (Woman FGD Participant).
>
> *“To be honest, it is important for these mothers to be empowered to buy water and support the strong fences, otherwise it will be very difficult to have good and sustainable gardens, even the few households you found have maintained growing home vegetable gardens most of them have good economic status to buy water, wells, fence or live close to water sources”* (Male IDI Participant).

All participants reported water for gardening to be a challenge in terms of availability, access and cost. It was informed that water availability was not a big problem in nearly all study villages because there are enough public and private water taps and wells. However, the cost of water was a big constraint as most participants could not afford the cost of water for domestic and home garden uses.

> *“Cost for water is the biggest challenge let me tell you, because you have to buy water for cooking, drinking, washing clothes, and bathing these are compulsory and requires money; so you choose not to irrigate vegetables until I get extra money”* (Woman FGD participant).
>
> *“One of the biggest challenge is the problem of access to water and its costs. Therefore, farmers had to seek water from those with wells and water pipes where the water was sold at a highest cost compared to their economic status. Even in the rainy season water was sold but the farmer did not have to buy water every day as sometimes it rained”* (Male IDI Participant).

Pests and diseases were reported to be a big problem even before the phase-out period of the homestead agriculture nutrition project in Rufiji, Tanzania considering that participants were discouraged from using industrial chemical pesticides for safety reasons.

> *“pest and diseases was another problem, you can plant vegetables and grow very well but when it reaches a height of about five leaves, pests invade and eat the whole garden. It’s sad, you can even cry considering that I have spent time and cost for buying water”* (Woman FGD participant).
>
> *“Despite providing education to farmers to use natural remedies but pests and diseases were very high so the effectiveness of these natural remedies was limited, for example they used ashes, hot peppers, and neem leaves solution. These home remedies reduced the rate of pests and diseases by about 20% or 30% compared to industrial pesticides”* (Male IDI Participant).

Most of the participants in FGDs and IDIs reported a shortage of seeds as a constraint on the better sustainability of homestead vegetable gardening. They reported that vegetable seeds were provided timely and free of charge during the intervention phase as the project incentives.

> *“Before the phase-out of the intervention all seeds and manure or fertilizers were provided for free but now days I have to buy seeds and fertilizer from shops. Since Seeds and fertilizer are sold at high price sometimes it is difficult to afford”* (Woman FGD participant).

Majority of participants reported limited availability and accessibility of manure and fertilizer was affecting vegetable productivity because of poor soil structure and fertility.

> *“Our land is sand soil, so without fertilizer you cannot get anything. So when you grow vegetables you must apply manure or other fertilizers such as UREA so that the soil is improved for vegetables”* (Woman FGD participant).
>
> *“…*..*our land is not fertile enough, so to get good vegetables you have to look for animal manure or buy fertilizer from the shop. If you plant without fertilizer the seeds will germinate but will not grow well”* (Woman FGD participant).

It was reported that existing community and system factors such as lack of enough knowledge on the importance of a balance diet and absence of by-laws or effective policies to support vegetable production had affected homestead vegetable gardening sustainability. This has contributed to participants not having affordable agriculture inputs and the absence of effective field visit support from the extension worker.

> *“Actually there should be a direct government support on homestead vegetable community gardening including financial support or subsidy for agriculture inputs that will help the community to afford managing home gardening”* (Male IDI Participant).
>
> *“You know, before project phase-out agriculture extension workers were provided with fuel allowance for field household visits to provide technical support to project participants to have productive home gardens. But the fuel allowance ceased when the project phased-out and hence it’s hard for Extetion Workers or Community Health Workers to volunteer on reaching out to participants”* (Male IDI Participant).

## Discussion

The study findings showed a low level of participation in homestead vegetable gardening practices one-year after the phase-out of the intervention; approximately 20% of households sustained active homestead vegetable production. Shortage of seeds, organic manure and fertilizers were significant factors associated with failure to sustain homestead vegetable production. It was found in the discussion that, the high cost of water, pests and diseases control challenges, destruction from free-grazing animals, dry weather conditions, poor soil fertility, and limited capital to purchase agriculture inputs also influenced the low proportion of households to sustain the vegetable production.

Shortage of seeds was a significant factor influencing homestead vegetable sustainability. Participants who could afford seeds cost for their homestead garden after the phase-out of the intervention were more likely to have active homestead gardens throughout the year. The present study finding is consistent with the study conducted in Sri Lanka that reported a shortage of agricultural inputs such as seeds was a constraint for vegetable garden sustainability (11). Also, the intervention conducted by Helen Keller International in Bangladesh reported that the shortage of seeds was a limitation for homestead vegetable sustainability, especially when there was no regular supply of seeds (24). Self-reliance on agriculture inputs particularly seeds, may be the main factor for a sensitized household to practice homestead vegetable gardening.

Limited supply of manure and fertilizer was a significant factor influencing the sustainability of homestead vegetable production in the study area. Participants with the capacity to access or buy farmyard manure or fertilizer were more likely to have active gardens. This finding supports previous study conducted in three villages in Nkokobe Municipal in South Africa that reported shortage of fertilizer led to low homestead vegetable harvest and sustainability (25). High yield of vegetable production following the free supply of manure and fertilizers during the intervention phase of the study was not enough to empower participants to explore cheap and reliable sustainable sources of manure after the phase-out of the intervention. Skills imparted to participants on the production of compost manure and poultry farming as a source of manure was unsuccessful. Cheap and readily available manure sources and fertilizer will always play a great role in feeding crops such as vegetables in poor rural communities, as demonstrated in Southern Ethiopia (26).

Typically homestead vegetable gardens are rain-fed; however, irrigation is important when the rain season ends. The study highlights the high cost of water due to its shortage was one of the factors influencing homestead vegetable gardening practices. This also agreed with previous studies in South Africa and Sri Lanka (11,17,27) that reported water shortage as the limiting factor for the sustainability of homestead gardens. This could explain why there was low percentage of participation in homestead vegetable gardening among respondents because this was attributed to shortage of water (11).

Pests and diseases reported attacking homestead vegetable gardens affect homestead vegetable production. Participants relied on the use of biological means such as home remedies (hot peppers, ashes, and neems) to control pests and diseases to ensure safety to the consumers. However, using biological measures alone to control pests and diseases has proved to be less effective, especially in the tropical environment that favours agricultural resistant pests and diseases (28,29).

Failure to control free-grazing domestic animals, including goats, chickens and ducks, discouraged participants from sustaining vegetable gardening because these animals were eating their produce. Building a fence to protect the vegetables from animals was not feasible to most participants due to cost. Literature shows that free-grazing animals destroy homestead gardens due to a lack of strong fencing. For example, a study conducted in rural villages of South Africa showed 32.8% of participants reported garden destruction due to lack of fencing (17). Results perceived in this study might be due to the fact that the study community have adapted free-range grazing system that eventually affect home gardens.

This study results also report weather conditions as a predictive factor for the sustainability of homestead vegetable gardens. Most of the participants highlighted the likelihood of growing vegetables during the wet or rainy season instead of the dry season because of the high cost of water. This may explain low participation in vegetable gardening practices during a specific period in the year. The study conducted in Sri Lanka have also reported variability of weather affect homestead gardens sustainability (30).

Participants adopted some new strategies after the intervention phased-out, especially on the type and frequency of vegetables grown, aiming to overcome some gardening challenges. These include growing cassava, sweet potato, pumpkin and *moringa*. Easy access to seeds/seedlings, drought tolerance and low susceptibility to pests and diseases were the main driving factor for the choice of these vegetables. These findings are in agreement with a study conducted in rural villages of South Africa that determined challenges affecting home vegetable gardening 10 years after intervention phase-out. This could explain why participants who adopted resistance and traditional vegetables managed to continue engaging in home garden activities despite the challenging agriculture environment (17).

This study had potential limitations that may have impacted the findings and conclusions. First, the dataset did not include a variable on the community culture of the study respondents in the area. Consequently, contribution of culture to the sustainability of home vegetable gardening was not possible. Therefore, data could support a better understanding of the variable in relation to home gardens on how homestead vegetable gardening is culturally accepted. Second, the study covered only one district of the Coastal region; therefore, it is difficult to generalize the findings to the rest of the region. Hence, this study has good internal validity but may have weak external validity. Third, this study was cross-sectional so that the findings would be restricted to the area investigated during research execution.

To the best of our knowledge, this is the first study in Tanzania to comprehensively explore factors associated with the sustainability of homestead vegetable production, specifically one-year after the phase-out period of a donor-funded homestead agricultural nutrition-sensitive intervention. Existing individual, community and system challenges played a significant role to influence the sustainability of this type of intervention. Effective community sensitization and government support are necessary to ensure uptake and sustainability of agriculture nutrition-sensitive interventions.

## Data Availability

All data supporting the findings is contained within the manuscript. The dataset used during the study is available from the dryad digital repository at https://doi.org/10.5061/dryad.nzs7h44tq.

## Acknowledgement

The author would like to thank study participants in Rufiji district for their voluntary participation in this study. The author acknowledges the contribution of Selemani Mmbaga, Lameck Pashet, Naomi Urio, and Master of Science in Public Health Research (MSc-PHR) students at Nelson Mandela African Institution of Science and Technology, Arusha, and Bagamoyo campuses, Tanzania.

## Author’s contributions

Conceptualisation of the research idea, KM, DM; Study design, KM, DM, AM, ALB, HM, WF, JK, CRC, MB, EK; Field research, KM; Research supervison, DM, AM, EK; Statistical analysis, Synthesis and interpretation, KM, DM, HM; Origianl manuscript draft, KM; Critical manuscript review, DM, AM, EK, ALB, HM, WF, JK, RAN, CRC, MB; All authors reviewed the results and approved the final version of the manuscript.

